# Silver diamine fluoride and oral health-related quality of life: A network meta-analysis

**DOI:** 10.1101/2021.04.05.21254786

**Authors:** Ryan Richard Ruff, Rachel Whittemore, Martyna Grochecki, Jillian Bateson, Tamarinda Barry-Godin

## Abstract

**Objective:** Silver diamine fluoride (SDF) is an effective non-surgical treatment for dental caries which may also impact oral health-related quality of life (OHRQoL). The objective of this study was to conduct a network meta-analysis of SDF versus other standard of care therapies on OHRQoL.

**Data Sources:** Studies published in PubMed/MEDLINE, Scopus, or Web of Science through March 2021 with no date or language restrictions.

**Study Selection:** Any randomized controlled trial, cohort, or case-control study that included silver diamine fluoride as either a single or combinative treatment for dental caries and a quantitatively measured outcome for oral health-related quality of life was included.

**Data Extraction and Synthesis:** Potentially eligible studies were screened by two independent reviewers trained in conducting systematic reviews. Studies meeting inclusion criteria underwent a full-text review with data being extracted using a standardized form, including publication details, study methodology, outcomes, assessors, and sample information. Studies underwent a risk of bias assessment and quality of evidence evaluation. Quantitative synthesis was performed using network meta-analysis.

**Main Outcome(s) and Measure(s):** Oral health-related quality of life.

**Results:** 19 articles were returned following search strategies. Following screening, ten studies were evaluated for full-text eligibility and five were retained for meta-analyses. Studies included in quantitative synthesis were classified as a high degree of evidence, suggesting estimated effects are similar to true effects. Direct and indirect estimates from network meta-analysis indicated that OHRQoL in children was not significantly different when treated with SDF versus atraumatic restorations (d = 0.02, 95% CI = -0.32, 0.36) or placebo (d = 0.03, 95% CI = -0.16, 0.22).

**Conclusions:** Evidence from the literature consistently shows no discernible impact on OHRQoL across various non-surgical treatments for dental caries. Overall oral health-related quality of life may increase regardless of treatment protocol due to treatment of the underlying disease. Concerns over the staining of dental decay and oral mucosa resulting from treatment with silver diamine fluoride do not seem to affect OHRQoL.

## 1. Introduction

Silver diamine fluoride is a novel therapy for the non-surgical treatment and prevention of dental caries, primarily delivered as a 38% concentration solution consisting of 24-27% silver, 7.5-11% ammonia, and 5-6% fluoride (Crystal and Niederman, 2016). Systematic reviews demonstrate that SDF is highly effective at arresting dental caries (Schmoeckel et al., 2020; Trieu et al., 2019). The comparative simplicity and efficiency of applying SDF make it an attractive alternative to traditional nonrestorative treatments (Gooch et al., 2009; Griffin et al., 2016), commonly used in community settings to mitigate the substantial burden of disease in underserved populations (Contreras et al., 2017). When applied to dental decay, the oxidizing effects of silver diamine fluoride results in irreversible black stains and superficial staining of the oral mucosa, potentially leading to aesthetic problems and negative impacts on oral health-related quality of life (Crystal et al., 2017a; Kyoon-Achan et al., 2020).

Oral health-related quality of life (OHRQoL) is a multidimensional construct consisting of subjective evaluations of oral health, functional well-being, emotional well-being, satisfaction with care, and sense of self (Ruff et al., 2017). Prior research suggests that dental caries negatively affects oral health-related quality of life (Chaffee et al., 2017). The focus on socio-psychological and cultural outcomes related to QoL, in addition to more traditional biological change, encourages greater consideration of orofacial appearance and overall aesthetics in the treatment of oral diseases. It is therefore possible that prototypical quality of life may increase due to a reduction of the burden of disease, yet simultaneously harm subjective perceptions of self (Sischo and Broder, 2011).

Research on the secondary effects of treating dental caries with silver diamine fluoride on quality of life yields mixed results, with SDF treatment demonstrating both improvements and no discernable effect on OHRQoL in children (Duangthip et al., 2019; Jiang et al., 2020, 2019; Rodrigues et al., 2020; Sihra et al., 2020). Similarly, the difference in OHRQoL when comparing SDF to atraumatic restorative treatment has previously shown to be both significant and non-significant (Rodrigues et al., 2020; Vollu et al., 2019). These results are further complicated in that silver diamine fluoride is occasionally applied in different populations as a combinative treatment with other preventive therapies for caries, such as fluoride varnish or dental sealants (Ruff and Niederman, 2018a). The objective of this study was to assess the comparative effects of SDF on OHRQoL relative to other therapies using network meta-analysis.

## 2. Methods

### 2.1. Search and extraction

This study is reported using the PRISMA checklist for network meta-analyses (PRISMA-NMA) (Hutton et al., 2015). The MEDLINE/PubMed, Web of Science, and Scopus databases were used with no restriction on language or date of publication. A search of grey literature was not performed. The last search was performed on 8 March, 2021. The search strategy was developed following the PICO question “What is the effect of silver diamine fluoride versus other surgical or non-surgical interventions for dental caries on oral health-related quality of life in subjects of any age?” We included any randomized controlled trial, cohort, or case-control study that included silver diamine fluoride as either a single or combinative treatment for dental caries and a quantitatively measured outcome for oral health-related quality of life. The complete search strategy was as follows: ((((silver diamine fluoride) OR sdf) OR diamine silver fluoride) OR silver ammonia fluoride) AND (((((((dental caries[MeSH Terms]) OR dental caries) OR caries) OR tooth decay) OR dental decay) OR carious lesion) OR dmf) AND ((((quality of life) OR qol) OR oral health related quality of life) OR ohrqol). The review was not registered in PROSPERO.

Potentially eligible studies were first independently screened by two reviewers (TBG and RW) who were previously trained for systematic reviews. Eligible studies were those that met study inclusion criteria: studies must have used silver diamine fluoride for the treatment of dental caries and must use a validated quantitative instrument for oral health-related quality of life. Any study that lacked a control group or comparator was excluded from quantitative synthesis but not from qualitative review. All comparative interventions were included, such as atraumatic restorations (ART), glass ionomer sealants, traditional amalgam restorations, fluoride varnish, or combinations of therapies (e.g., ART plus fluoride varnish). Studies that met inclusion criteria underwent a full-text review with the following data being extracted using a standardized form: publication details (authors, title, and year of publication), study methodology (design type, treatment, comparator), outcome measure (e.g., COHIP, ECOHIS) and assessor (parent or child), and sample information (sample size, treatment effect, and standard error/standard deviation).

### 2.2. Risk of bias assessment

Risk of bias of included studies was independently evaluated (RW and TBG) using the Newcastle-Ottawa Scale (NOS) for assessing the quality of non-randomized studies (Wells et al., 2021) and the revised Cochrane risk-of-bias tool for randomized trials (RoB 2) (Higgins and Green, 2008; Sterne et al., 2019). Reviewers ranked each item included in NOS and RoB 2 forms as low risk of bias, high risk of bias, or unable to identify. Disagreements were resolved via a third reviewer (RRR).

### 2.3. Quality of evidence evaluation (GRADE)

The GRADE tool was used to asses quality of evidence of studies included in network meta-analyses (Higgins and Green, 2008). Each study was evaluated and assigned a quality score of very low (true effect probably markedly different from estimated effect), low (true effects might be different), moderate (true effects probably close to estimated effects), and high (a great deal of confidence that true effects are similar to estimated effects).

### 2.4. Data Synthesis

Overall pooled analysis of direct evidence was computed using a random effects meta-analysis, which also produced consistent treatment effects and standard errors for each study. Heterogeneity was determined using the Q and I^2^ statistics. Direct and indirect comparisons across individual treatments across all studies were then evaluated using a frequentist network meta-analysis. Mean differences and standard errors were included for each study. Network geometry was evaluated and it was determined that two subnetwork analyses were required due to node disconnection. Individual network meta-analyses were then performed on each subnetwork and network graphs were computed. Node split analyses were not performed due to the small number of studies included in analysis. Treatment ranking from the network meta-analysis was calculated using the P-Score ranking metric, analogous to the Surface Under Cumulative RAnking (SUCRA) method. Tables for direct and indirect evidence as well as forest plots or subnetworks were computed. No studies included had multiple arms. All treatments included in NMA were performed in similar subject populations, suggesting the networks met the transitivity assumption. Analysis was performed using R v4.0.2.

## 3. Results

Nineteen articles were returned after searching. There were no duplicate records. Nine articles were excluded following initial screening yielding ten full-text records assessed for eligibility Ruff et al. (2021). Five of these studies were excluded, all due to a lack of a control group or adequate comparator (e.g., single sample pre-post designs). Five studies were therefore included in quantitative synthesis (Figure 1). Major study characteristics of studies included in network meta-analyses (Table 1) show that all included articles were randomized controlled trials; all of the articles reviewed for full-text eligibility that were excluded were either cross-sectional or prospective cohort studies. Among the included studies, four assessed oral health-related quality of life using either the child or parent form of the Early Childhood Oral Health Impact Scale (ECOHIS) and one used the Child Oral Health Impact Profile (COHIP). Treatments consisted of SDF or SDF+FV, while comparators included placebo, ART, and ART+FV. The average sample size across all studies was 126.

**Table 1:** Study characteristics

| Author/Year | Country | Design | OHRQoL | Assessor | Ages | Treatment | Comparator | N | NMA? |
| --- | --- | --- | --- | --- | --- | --- | --- | --- | --- |
| Jiang et al, 2020a | China | RCT | C-ECOHIS | Parent | 3-4 y | SDF | Placebo | 253 | Yes |
| Cernigliaro et al, 2019 | USA | Cross-sectional | ECOHIS | Parent | 0-14 y | SDF | N/A | 48 | No |
| Duangthip et al, 2018 | China | Cohort | C-ECOHIS | Parent | 4-5 y | SDF | N/A | 226 | No |
| Ruff et al, 2021 | USA | RCT | COHIP-SF | Child | 5-13 y | SDF+FV | ART+FV | 246 | Yes |
| Hiremath et al, 2020 | India | Cross-sectional | COHIP-SF | Child | 12-16 y | SDF | N/A | 84 | No |
| Jiang et al, 2019 | China | RCT | C-ECOHIS | Child | 3-4 y | SDF | Placebo | 187 | Yes |
| Rodrigues et al, 2020 | Brazil | RCT | B-ECOHIS | Parent | 2-5 y | SDF | ART | 108 | Yes |
| Shira et al, 2020 | Canada | Cohort | ECOHIS | Parent | 0-6 y | SDF+FV | N/A | 40 | No |
| Vollu et al, 2019 | Brazil | RCT | B-ECOHIS | Parent | 3.62 y | SDF | ART | 26 | Yes |
| Renugalakshmi et al, 2021 | Saudi Arabia | Cohort | A-ECOHIS | Parent | 2-6 y | SDF | N/A | 51 | No |

**Figure 1:**
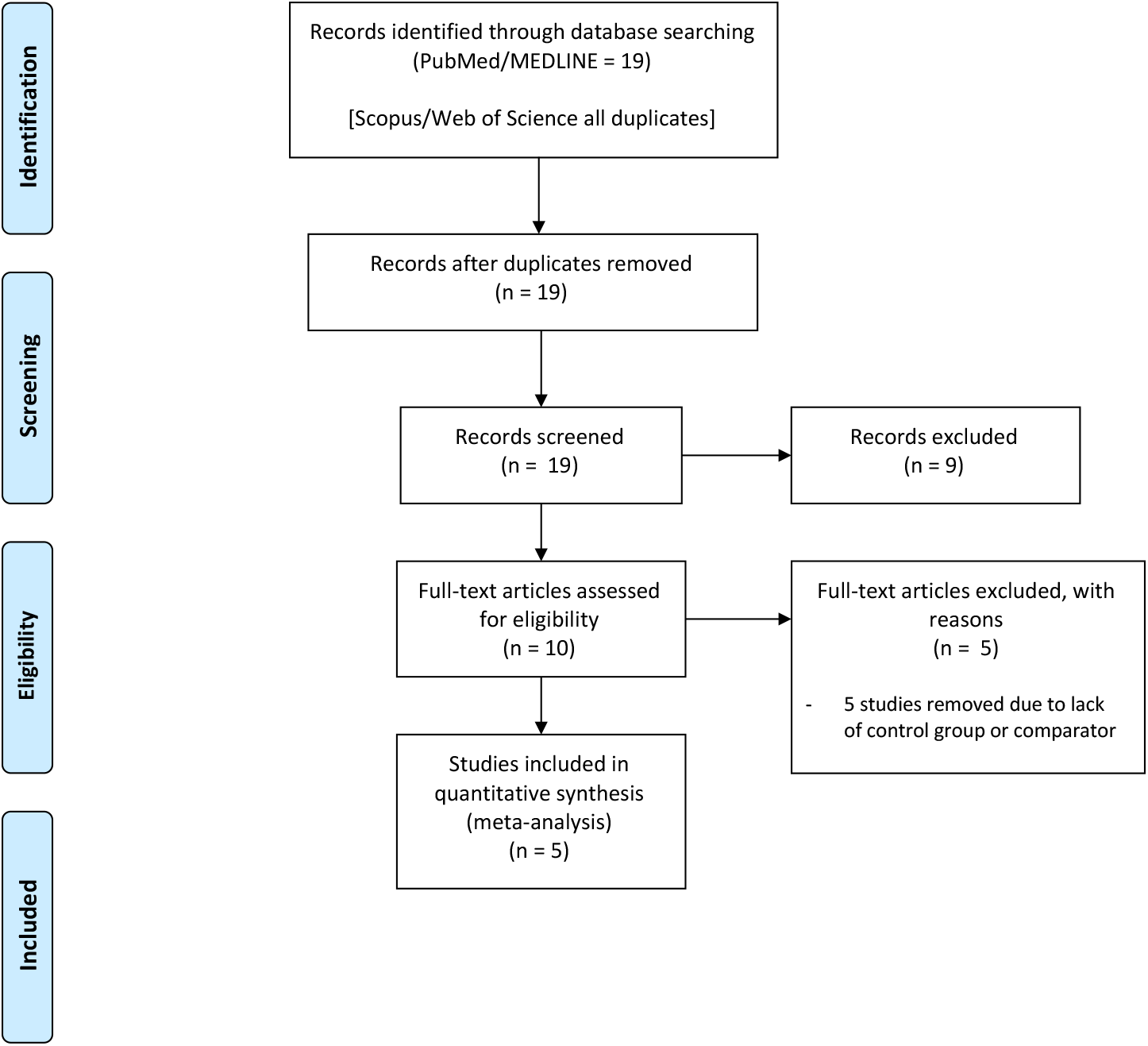
PRISMA Diagram

### 3.1. Risk of bias

Bias assessment for randomized studies included in quantitative synthesis (Table 2) indicated that three of the four studies had some concerns of bias due to the likelihood that both participants and caregivers were aware of the assigned intervention (Jiang et al., 2020, 2019; Vollu et al., 2019). Specifically, the likelihood of participants experiencing the staining side effect characteristic of silver diamine fluoride, despite the presence or absence of patient and operator blinding, may have contributed to deviations from the intended intervention. However, none of the studies provided information to this effect. A single study had a high risk of bias due to concerns regarding allocation concealment and missing outcome data for approximately 9% of participants (Rodrigues et al., 2020). In contrast, non-randomized cohort and case-control studies not included in meta-analyses (Table 3) all included a clinical examination for exposure assessment, had relatively lengthy follow-up periods, and exhibited 100% retention of study participants. However, all studies used convenience sampling with no randomization and lacked an adequate comparator. Finally, two cross-sectional studies evaluated (Table 4) used appropriate statistical methods but similarly lacked an acceptable comparator, did not justify the sample size, and provided no information on the characteristics of non-responders.

**Table 2:**
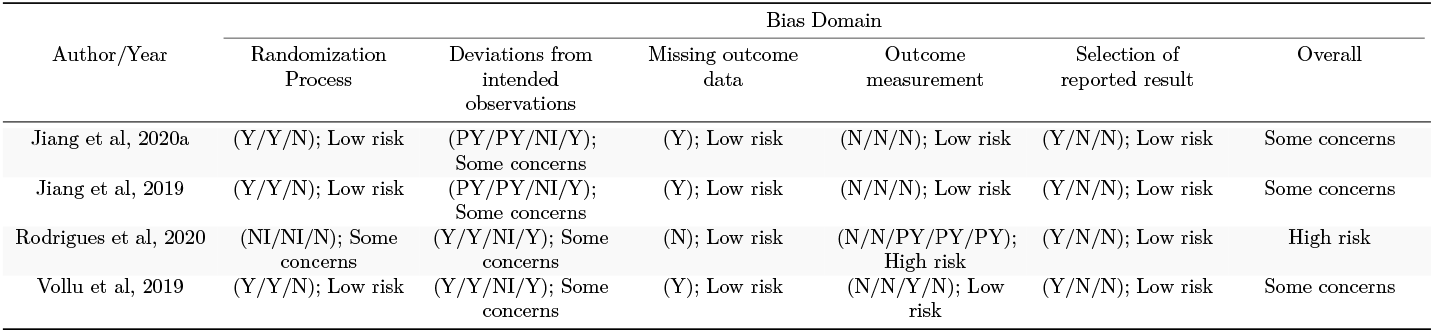
Risk of bias assessment for randomized studies. (Y=yes, N=no, referring to each category included on the RoB2 assessment tool)

| Author/Year | Bias Domain |  |  |  |  | Overall |
| --- | --- | --- | --- | --- | --- | --- |
|  | Randomization Process | Deviations from intended observations | Missing outcome data | Outcome measurement | Selection of reported result |  |
| Jiang et al, 2020a | (Y/Y/N); Low risk | (PY/PY/NI/Y); Some concerns | (Y); Low risk | (N/N/N); Low risk | (Y/N/N); Low risk | Some concerns |
| Jiang et al, 2019 | (Y/Y/N); Low risk | (PY/PY/NI/Y); Some concerns | (Y); Low risk | (N/N/N); Low risk | (Y/N/N); Low risk | Some concerns |
| Rodrigues et al, 2020 | (NI/NI/N); Some concerns | (Y/Y/NI/Y); Some concerns | (N); Low risk | (N/N/PY/PY/PY); High risk | (Y/N/N); Low risk | High risk |
| Vollu et al, 2019 | (Y/Y/N); Low risk | (Y/Y/NI/Y); Some concerns | (Y); Low risk | (N/N/Y/N); Low risk | (Y/N/N); Low risk | Some concerns |

**Table 3:**
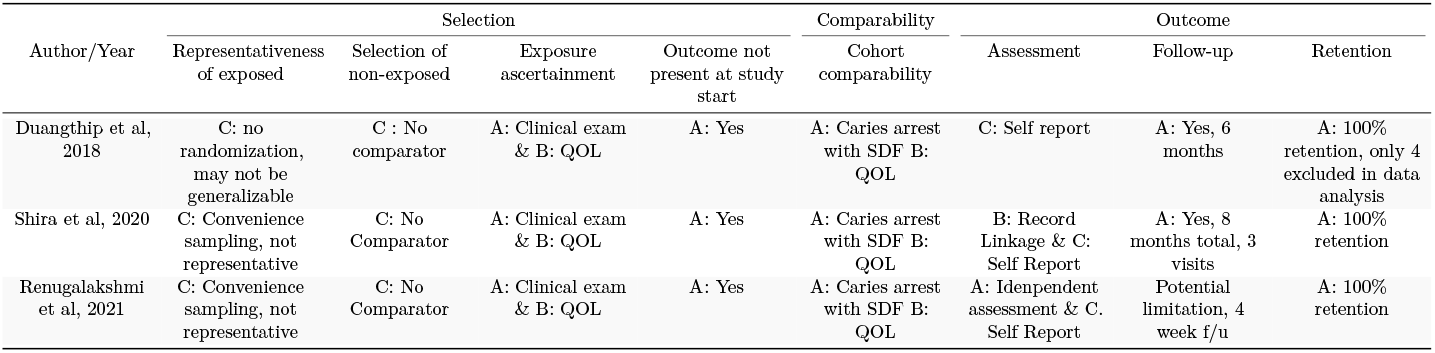
Risk of bias assessment for non-randomized studies (A, B, and C refer to coding as specified in the NOS manual)

**Table 4:** Risk of bias assessment for cross-sectional studies (A, B, and C refer to coding as specified in the NOS manual)

| Author/Year | Selection |  |  |  | Comparability | Outcome |  |
| --- | --- | --- | --- | --- | --- | --- | --- |
|  | Representativeness of exposed | Sample Size | Non-Respondents | Risk Factor | Comparability | Assessment | Statistical Test |
| Cernigliaro et al., 2019 | C: Selected group of users | B: Not justified | C: No description or non-response rate | B: Measurement tool is described | n/a | C: Self report | A: Appropriate |
| Hiremath et al., 2020 | C: Selected group of users | B: Not justified | C: No description or non-response rate | B: Measurement tool is described | n/a | C: Self report | A: Appropriate |

### 3.2. Quality of evidence

GRADE evaluation of retained studies (Table 5) was determined to be of a high degree of certainty, and effects pooled by meta-analyses may be considered similar to the true effect of SDF on OHRQoL. All included studies in NMA were randomized controlled trials with sufficient sample sizes, with high precision and consistency in effects across studies. Point estimates were similar and all confidence intervals overlapped, with no evidence of heterogeneity in results. Studies were conducted in different populations using established methods for assessing oral health-related quality of life (e.g., ECOHIS and COHIP) and utilized standards of care protocols for treatment.

**Table 5:** GRADE evaluation of quality of evidence

| Domain | Confidence | Reason |
| --- | --- | --- |
| OHRQoL | High | Low risk of bias; consistency of effects; good precision |

### 3.3. Meta-analysis

Meta-analysis results indicate that, irrespective of comparator, there was no significant difference in quality of life versus silver diamine fluoride. There was no heterogeneity among the included studies (I^2^ = 0.0%, Q = 0.75, p = 0.95, Figure 2). Pooled effects showed a difference in oral health-related quality of life that was not significantly different from zero (SMD = -0.06, 95% CI = -0.20, 0.08). Direct and indirect evidence from network meta-analyses show similarity in effects regardless of comparator used: there were no differences in OHRQoL between children treated with SDF versus placebo (MD = 0.03, 95% CI = -0.16, 0.22) and atraumatic restorative treatments (MD = 0.02, 95% CI = -0.32, 0.36); nor were there any differences between ART and placebo (MD = -0.01, 95% CI = -0.40, 0.38) or between SDF+FV versus ART+FV (MD = 0.16, 95% CI = -0.1, 0.42). Effects from network meta-analyses are shown via forest plots (Figures 3 & 4).

**Figure 2:**
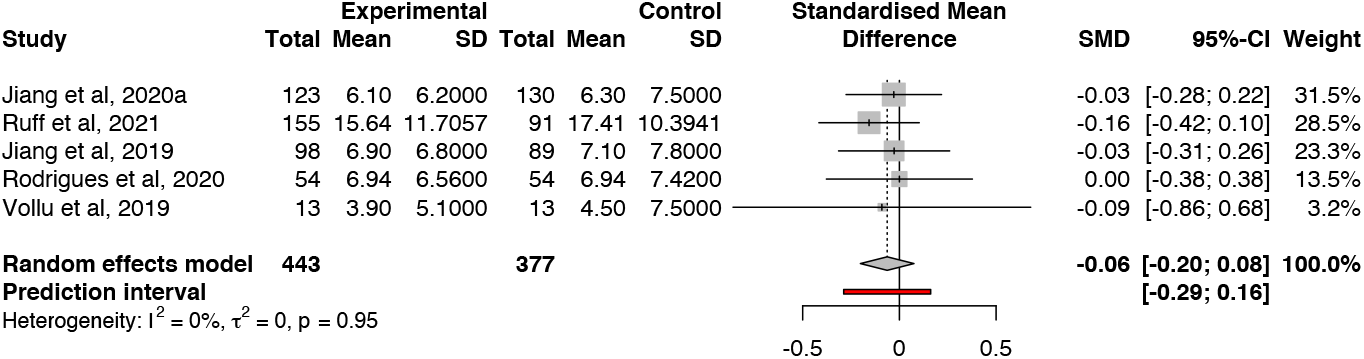
Random effects meta-analysis

**Figure 3:**
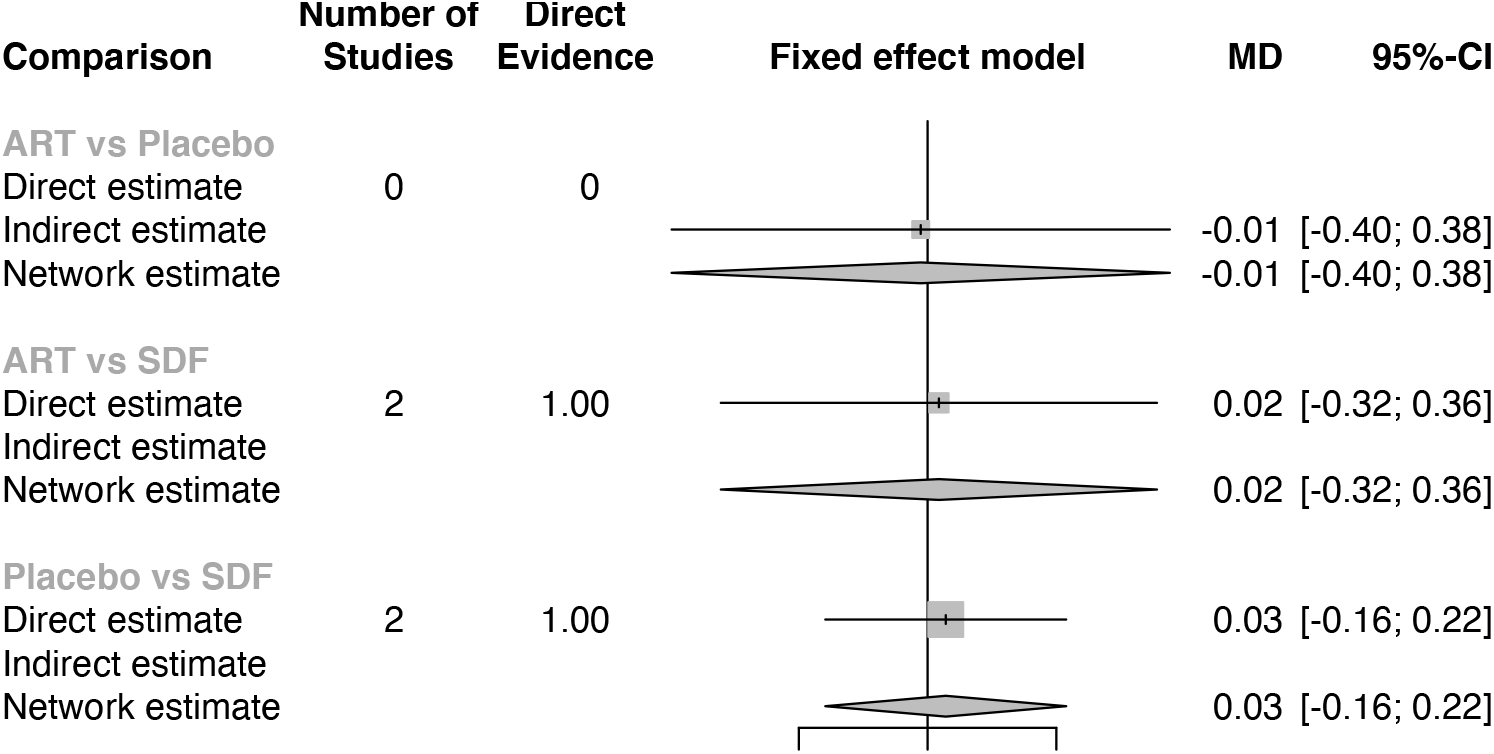
Subnetwork 1 forest plot, network meta-analysis

**Figure 4:**
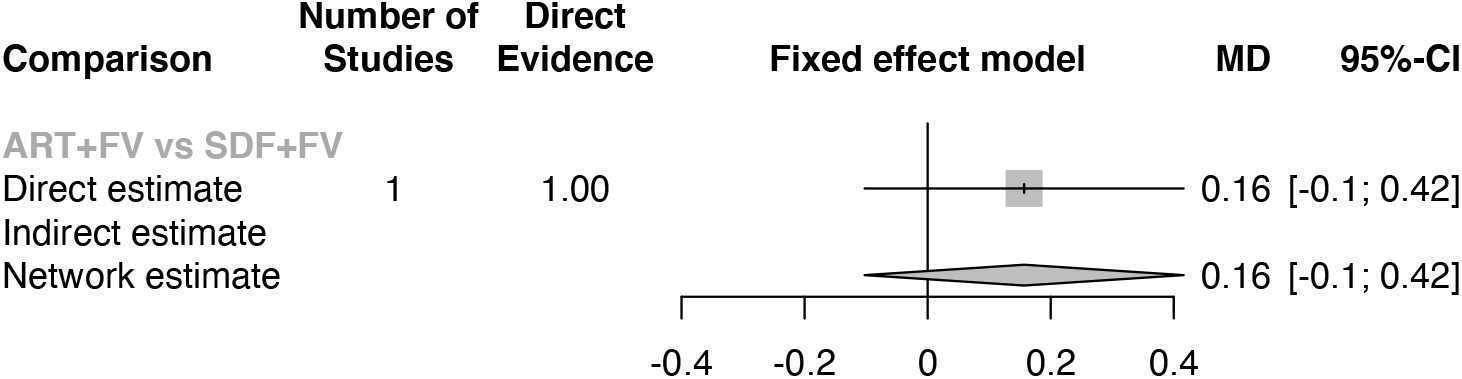
Subnetwork 2 forest plot, network meta-analysis

Results from treatment ranking indicate that the standard placebo had the highest P-score (0.5689), followed by ART (0.5095) and SDF (0.4216). In this context, P-scores are interpreted as the extent that any treatment is better than any other treatment. Similarity in P-scores in these results indicate comparability of treatment on OHRQoL.

## 4. Discussion

The American Association of Pediatric Dentistry supports the use of silver diamine fluoride as part of a caries management plan and provides clinical practice guidelines for its use (Crystal et al., 2017b). Single-use applications in pediatric populations for caries arrest has demonstrated effectiveness ranging from 47-90%, and SDF-arrested lesions can either be later restored as part of traditional surgical caries treatment or perpetually reinforced with annual or bi-annual reapplications. The off-label use of SDF is particularly attractive for high-risk populations as an effective, efficient non-surgical therapy for untreated caries (Ruff and Niederman, 2018a, 2018b). Previous research on the acceptability of silver diamine fluoride suggests that the staining effect of arrested lesions in primary teeth is more tolerated by parents when applied to posterior teeth than in anterior teeth. Notably, concerns over teeth staining from SDF depended on the presence of extant behavioral issues of the child towards dental care or whether alternative treatments for unmet disease required more invasive measures such as general anesthesia (Seifo et al., 2020).

Facial aesthetics are a potential significant influence on perceptions of self, such as in children with orofacial anomalies (Ruff et al., 2016) and in adolescents seeking orthodontic treatment (Phillips and Beal, 2009). In particular, self-perceptions of appearance and positive feelings of dentofacial regions were related to self-concept (Phillips and Beal, 2009). Personal beliefs of self-concept may be related to health-related quality of life, and suitable QoL measures are those that include ideographic and subjective approaches to self-concept, such as values, feelings, experiences, and attitudes towards self in the contest of relationships and the world in general (Zlatanović, 2000). This connection emphasizes the need to explore any unintended consequences of the use of treatments for oral disease that might negatively impact facial appearance and sense of self.

Our findings indicate that while there is no comparative difference in OHRQoL among children receiving silver diamine fluoride versus other standard of care treatments such as atraumatic restorations, excluded studies suggest that there may be general improvement in OHRQoL over time due to treating underlying disease. Studies that only assessed within-subject change in oral health-related quality of life prior to and after treatment with silver diamine fluoride for dental caries were not quantitatively evaluated in this study due to a lack of an adequate comparator. Of these excluded studies, two found that OHRQoL/caregiver satisfaction improved (Cernigliaro et al., 2019; Renugalakshmi et al., 2021), two showed no appreciable change (Duangthip et al., 2019; Sihra et al., 2020), and one showed a negative effect on OHRQoL (Hiremath et al., 2020), though this latter study used inappropriate statistical analysis for a single-sample repeated measures design. Longitudinal research on the change in ORHQoL both within SDF treatment and compared to other treatments would support a greater understanding of the long-term impact on quality of life.

The small number of clinical trials of silver diamine fluoride that include measures for subjective quality of life prohibit analyses by severity of disease. We were therefore unable to explore whether the baseline severity of untreated caries treated by SDF had any impact on OHRQoL. Some studies have shown that oral health-related quality of life was negatively impacted by the general increase in severity (Corrêa-Faria et al., 2018; Fernandes et al., 2017), therefore it may be that the negligible impact of SDF on OHRQoL relative to other interventions is relevant only at certain levels of disease burden. Similarly, included studies did not stratify by whether SDF was applied on posterior versus anterior teeth. Indeed, some pragmatic studies of SDF did not include anterior teeth application in their clinical protocols (Ruff and Niederman, 2018a, 2018b).

The diversity of available non-surgical therapies for dental caries (e.g., atraumatic restorative treatments, fluoride varnish or gels, glass ionomer sealants, or combinations of these interventions) means that numerous studies are necessary for a fully connected network. The disconnected networks presented in this analysis due to inadequate support therefore limits estimates of effects, specifically the disconnection for packaged treatments of SDF plus FV versus ITRs plus sealants. While we considered incorporating nonrandomized studies in analysis to expand upon the network, the lack of a comparator in these studies jeopardized the plausibility of the transitivity assumption (Rouse et al., 2016). Further studies that can improve the connectivity of the network by providing missing links between treatments is recommended.

Despite these limitations, our study suggests that overall oral health-related quality of life is not appreciably affected by silver diamine fluoride treatment for dental caries when compared to other standard of care interventions and results are strengthened by the similarity of effects across alternative treatments. The impact of the baseline severity of disease treated by SDF or the role of anterior versus posterior treatment on OHRQoL is still unknown.

## Data Availability

All data and code is available at a Github repository operated by the corresponding author.

https://github.com/ryanruff/SDF_QoL_NMA

## 5. Author Contribution

RRR conceived of the study, developed the search strategy, conducted the quality evaluation, and performed statistical analyses. TBG and RW performed abstract screening and risk of bias assessments. JB and MG performed full-text review and data extraction. All authors contributed to writing and revising the manuscript.

## 6. Conflict of Interest

The authors report no conflicts of interest.

## Notes

### Competing Interest Statement

The authors have declared no competing interest.

### Funding Statement

No external funding was received.

